# Role of Cardiac Rehabilitation in Improving Outcomes after Myocardial Infarction

**DOI:** 10.1101/2023.10.20.23297313

**Authors:** Raghu Ram Shanmukh Nemani, Bala Sushritha Gade, Dedeepya Panchumarthi, Bhargava Venkata Sasidhar Reddy Bathula, Binay Kumar Panjiyar

**Author notes:** **First author:** Raghu Ram Shanmukh Nemani, MBBS [ ]. **Co-authors:** Bala Sushritha Gade, MBBS [ ], Dedeepya Panchumarthi, MBBS [ ], Sasidhar Reddy Bathula, MBBS [ ].

## Abstract

Myocardial Infarction (MI) an integral part of Acute Coronary Syndrome (ACS) occurs due to Atherosclerotic narrowing of the Coronary blood vessels. ACS being one of the major cardiovascular diseases (CVDs) has led to a significant amount of mortality & morbidity. The post-myocardial infarction period greatly impacted the physical; psychological; social; emotional; and occupational well-being. Cardiac rehabilitation (CR) helps by addressing these immediate effects & also improves long-term well-being and overall quality of life. The benefits of CR include enhanced exercise capacity, risk factor reduction, improved quality of life (QOL), and reduced mortality and hospital readmission. We used a systematic literature review (SLR) approach in his article to provide a global overview of the cutting edge offered by CR in the post-MI phase. We reviewed 33 articles from journals of good repute published between 2013 and August 15, 2023, focusing on six selected papers for in-depth analysis. The analysis was focused on factors such as positive outcomes of CR, and the effects of CR in post-myocardial infarction. Even though there is a lack of statistically significant evidence for the benefits of CR; Meta-analyses confirm its positive impact on cardiovascular outcomes, such as decreased mortality, cardiac events, and hospital Readmissions. There is still a need for ongoing research to enhance the understanding of its mechanisms and statistically prove its effectiveness. As CR continues to evolve, referral and participation in CR should be increased as it improves overall health and well-being.

## Introduction and Background

MI happens as a result of partial or complete blockade of blood flow (coronary) to the heart muscle, leading to ischemic death of the tissue supplied by that artery. Acute myocardial infarction (AMI) is classified into two types based on electrocardiogram (ECG) findings: ST-segment elevated myocardial infarction (STEMI) and non-ST-segment elevated myocardial infarction (NSTEMI) [1]. STEMI, occurring due to complete blockade, requires an emergent opening of the blocked vessel(s); Cardiologists emphasize a critical statement "The heart muscle is dependent on time, and time equates to life." [2]. But in NSTEMI there is no ticking clock since it’s a partial occlusion, which allows some flow of blood to the tissue

AMI being one of the most common CVDs, continues to pose a significant health burden globally. Approximately 605,000 new instances of myocardial infarction occur every year, along with 200,000 cases of recurring attacks. Within this total of 805,000 initial and repeated events, around 170,000 are believed to be asymptomatic [3]. The recent COVID-19 pandemic, still affecting populations due to its frequent emergence of new variants, significantly influenced cases of acute myocardial infarction (AMI). A few factors contributing to this were that, amidst the COVID-19 pandemic, timely interventions such as the Door-to-balloon time faced delays [4], and the occurrence of arterial thromboembolic events following COVID-19 contributed to a rise in the frequency of AMI [5].

Amid the high incidence and prevalence of AMI, there is an increasing need for effective interventions that not only address both immediate medical concerns and long-term health outcomes and quality of life. Cardiac rehabilitation (CR) is an emerging intervention targeting the above concerns. CR includes various actions and approaches that empower patients to take control of their condition, prevent disease progression, and potentially reverse its course [6]. CR is a critical component of cardiovascular care for patients with cardiovascular disease, entitling it to a Class 1a recommendation. It involves a structured program specific to each patient that typically lasts 3 to 4 weeks [6].

CR is a cost-effective strategy with a holistic approach, encircling physical, emotional, and social aspects of patients’ Health-Related Quality of Life (HRQOL) [7]. The primary aim of CR is speeding up the secondary prevention and improvement of patients’ quality of life (QOL). The core elements of CR comprise exercise training, lifestyle modification, and psychological intervention [6]. CR extends beyond mere exercise routines to encompass comprehensive patient care, including drug therapy optimization, nutritional guidance, smoking cessation, stress management, and lifestyle improvement [8]. This comprehensive approach targets various aspects of patient health, ranging from physical fitness and cardiovascular risk factors to mental well-being.

CR’s goals include enhancing exercise tolerance, optimising coronary risk factors (e.g., lipid profiles, blood pressure, glucose levels), and addressing psychological aspects such as stress, anxiety, and depression [6]. CR brings about numerous positive outcomes. These encompass enhancements in exercise capacity, muscle strength, heart-related risk factors, and overall quality of life. Additionally, CR contributes to a reduction in mortality for patients with acute coronary syndrome (ACS)[8]. These advantages of CR extend to the elderly as well. CR has been shown to improve various essential patient outcomes, including exercise capacity, control of cardiovascular risk factors, quality of life, hospital readmission rates, and mortality rates [6]. The meta-analysis done by Haigang ji, which included the group that underwent cardiac rehabilitation (CR) and the non-CR group, indicated a substantially lower hazard ratio; major adverse cardiac events (MACE); and recurrence rate of MI in the CR group, denoting a considerable benefit [8].

Individuals participating in CR witness a range of positive effects. These include increased exercise capacity, improved management of cardiovascular risk factors, a better quality of life, decreased rates of hospital readmissions, and lower mortality rates. Heran and his colleagues organised 47 different studies involving 10,794 patients who were randomly assigned to either CR or standard care. The results of their analysis revealed noticeable reductions in both overall mortality and cardiovascular mortality [6]. A retrospective cohort study done by Kureshi, and Faraz showed no statistically significant difference in general health status in patients who took part in CR and did not take part. However, the study stated that participation in CR yields favourable survival benefits [9]. Furthermore, comprehensive evaluations of studies consistently emphasize the decreased mortality risk linked to engaging in CR. This has resulted in influential clinical directives, such as those provided by the American Heart Association and American College of Cardiology, designating referral to CR as a class IA cardiac care [9].

## Review

### Methods

Focusing on the outcomes of CR in Post MI patients, our review followed guidelines for Preferred Reporting Items for Systematic Reviews and Meta-Analyses (PRISMA) for 2020 in Figure *1* and only used data collected from published papers, eliminating the need for ethical approval.

**Figure 1:**
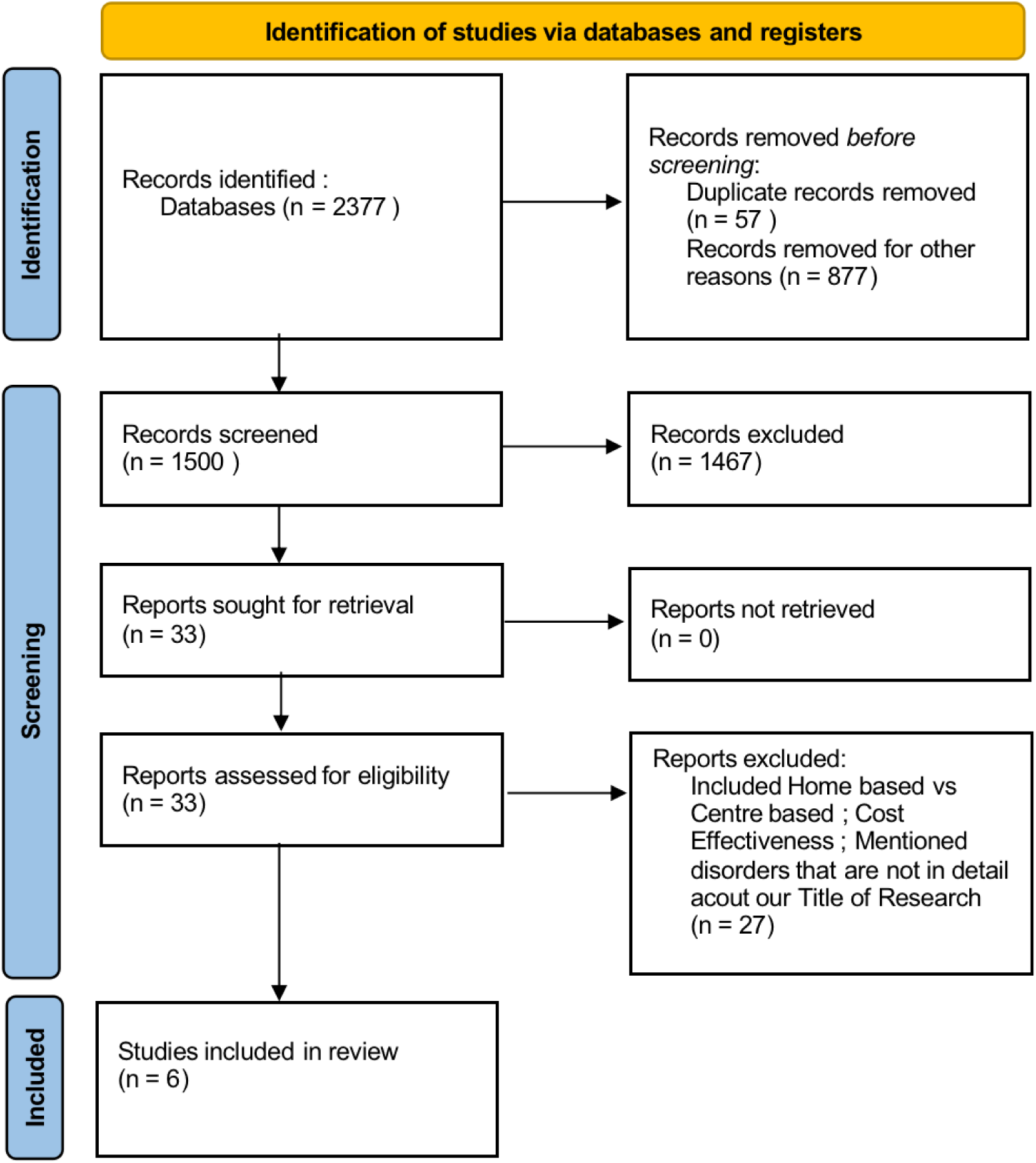
Flow diagram of PRISMA illustrating the search strategy and study selection process for the systematic review.

PRISMA:- Preferred Reporting Items for Systematic Reviews and Meta-Analyses

### Systematic Literature Search and Study Selection

We conducted a thorough search for relevant publications by using PubMed, including Medline and Google Scholar. We searched for studies mentioned in review papers, editorials, and commentaries on PubMed. Nevertheless, we continued searching for additional studies that satisfied our inclusion criteria.

We had a list of abstracts that we independently reviewed for inclusion using specific criteria. The criteria included using Cardiac Rehabilitation, focusing on Outcomes in Post Myocardial Infarction. We excluded review papers and animal studies. reviewers conducted a dual review, and disagreements were resolved through discussion.

### Inclusion and Exclusion Criteria

We established specific criteria for including and excluding participants to achieve our study goals. Our Criteria can be summarised in Table *1*.

|  | Inclusion Criteria | Exclusion Criteria |
| --- | --- | --- |
| 1. | Human studies | Animal studies |
| 2. | From 2013 to 2023 | Only methodological studies explaining programming details |
| 3. | English Text | Non-English texts |
| 4. | Gender: All |  |
| 5. | Age: > 19 years of age | Age:<=19 years |
| 6. | Free papers | Papers that needed to be purchased |
|  |  | Studies involving clinical data other than cardiovascular diseases |

### Search Strategy

The population, intervention/condition, control/comparison, and outcome (PICO) criteria were utilized to conduct a thorough literature review. The search was conducted on databases such as PUBMED (including Medline) and Google Scholar Libraries, using relevant keywords, such as cardiac rehabilitation, post-myocardial ischemia, and positive outcomes. The medical subject heading (MeSH) approach for PubMed (including Medline) and Google Scholar, as detailed in Table *2*, was employed to develop a comprehensive search strategy.

**Table 2:** Showing search strategy, search engines and the search results displayed.

|  | Database | Search Strategy | Search Results |
| --- | --- | --- | --- |
| 1. | PubMed | Cardiac Rehabilitation or Post Myocardial Infarction | 176 |
|  |  | Positive Outcome and Cardiac Rehabilitation or Myocardial Infarction | 2076 |
| 2. | Google Scholar | Role of Cardiac Rehabilitation in Improving Outcomes after Myocardial Infarction<br>- Took relevant articles from the first 30 pages | 125 |

### Quality Appraisal

To ensure the reliability of our chosen papers, we utilized various quality assessment tools. We employed the PRISMA checklist and Cochrane bias tool assessment for randomized clinical trials for systematic reviews and meta-analyses. Non-randomized clinical trials were evaluated using the Newcastle-Ottawa tool scale. We assessed the quality of qualitative studies, as shown in Table *3*, using the critical appraisal skills program (CASP) checklist. To avoid any confusion in the classification, we utilized the scale for the assessment of narrative review articles (SANRA) to evaluate the article’s quality.

**Table 3:** Showing quality appraisal tools used.

| Quality Appraisal Tools Used | Type of Studies |
| --- | --- |
| Cochrane Bias Tool Assessment | Randomized Control Trials |
| Newcastle-Ottawa Tool | Non-RCT and Observational Studies |
| PRISMA Checklist | Systematic Reviews |
| SANRA Checklist | Any Other Without Clear Method Section |

PRISMA: Preferred reporting items for systematic reviews and meta-analyses ; SANRA: Scale for the assessment of non-systematic review articles

## Results

After searching through three selected databases, PubMed, Medline, and Google Scholar, we extracted 2377 articles. We removed 57 duplicate articles. We then carefully reviewed each paper and applied specific criteria, which led to the exclusion of 2170 articles. From the remaining 150 articles, we chose not to utilize 122 of them due to unsatisfactory titles and abstracts. We closely examined the remaining 28 articles and excluded 21 more as their content was not specific to our title of research. Finally, we conducted a thorough quality check on the remaining seven articles, which all met our criteria. These seven articles are included in our final systematic review. Table 4 provides a detailed description of each.

**Table 4:** Summary of the results of the selected papers.

| **Author/Year** | **Study Design** | **Database**<br>**Used** | **Article Name** |
| --- | --- | --- | --- |
| Tessler, Joseph /<br>2019 | Comprehensive<br>Review | Google Scholar | Cardiac rehabilitation |
| Bellmann,<br>Barbara / 2020 | Comprehensive<br>Review | Google Scholar | The beneficial effects of cardiac<br>rehabilitation |
| Kureshi Faraz,<br>Kevin. F / 2016 | Retrospective<br>Cohort Study | Google Scholar | Association between cardiac rehabilitation<br>participation and health Status Outcomes<br>after acute myocardial infarction |
| Troy Francis,<br>Nader Kabboul /<br>2019 | Meta-Analysis | Google Scholar | The effect of cardiac rehabilitation on health-related quality of life in patients with coronary artery disease: a meta-analysis |
| Haigang Ji, Liang<br>Fang / 2019 | Meta-Analysis | PubMed | Effects of Exercise-Based Cardiac Rehabilitation in Patients with Acute Coronary Syndrome: A Meta-Analysis |
| Richard Powell,<br>Gordon<br>McGregor / 2018 | Systematic Review<br>& Meta-Analysis | PubMed | Is exercise-based cardiac rehabilitation effective? A systematic review and meta-analysis to re-examine the evidence |

## Discussions

Cardiac rehabilitation consists of three phases [10].

1. Clinical phase: Starts after heart event/ completion of intervention, by assessing patient’s physical abilities. It includes guiding the patient through non-strenuous exercises in the bed or bedside.
2. Outpatient rehab: Begins post-stability and cleared by cardiology. It has a custom exercise plan tailored to specific patients based on assessments focusing on identifying limitations in physical function, and activities.
3. Post-rehab: Enhances flexibility, strength, and aerobic fitness. Patients are encouraged to maintain an active lifestyle. Regular outpatient check-ups are advised to monitor cardiovascular health and medication regimens.

The components of CR include patient assessment, nutritional counselling, weight management, blood pressure management, lipid management, diabetes management, tobacco cessation, psychosocial management, physical activity counselling, and exercise training.

We don’t completely understand the precise mechanism by which CR enhances prognosis for individuals with AMI [8]. However CR offers numerous physiological advantages due to its emphasis on physical activity. Participating in exercise training has been proven to enhance maximal oxygen uptake (VO2max), and enhance the flow of blood to the heart muscle [10]. It also leads to decreased smoking, body weight, serum lipids, and blood pressure [10]. As a part of CR, exercise training has been shown to enhance the endothelium-dependent dilation of coronary arteries in patients with ischemic heart disease [11]. CR has shown various direct benefits for the cardiovascular system. These encompass improvements in the supply of oxygen to the heart muscle, functioning of the endothelial layer, balance of the autonomic nervous system, factors related to blood clotting, markers of inflammation, and growth of collaterals for blood flow in the coronary arteries [8]. Because of these effects of CR, it has shown its positive impact on the patient.

CR has consistently demonstrated significant reductions in overall cardiovascular mortality and hospital admission rates. Studies conducted by Heran et al. and others have indicated a decrease in cardiovascular mortality and improved exercise capacity among patients participating in exercise-based CR [6]. A study by Richard Powell showed a reduction in hospital admission following CR which was even statistically significant [12]. A study conducted by Zalid A. Shah demonstrated that engaging in mild to moderate cardiac rehabilitation (CR) was linked to a lower likelihood of experiencing post-infarction angina and irregular heartbeats [13]. The above studies suggest that CR plays a crucial role in enhancing patient survival and reducing the likelihood of recurring cardiovascular events.

One of the noticeable advantages of CR is its positive impact on patients’ quality of life (QOL). Several studies have evaluated the effect of CR on QOL primarily using the SF 36 quality of life questionnaire, and their results consistently indicate that CR leads to significant improvements in various domains of QOL [14][15]. For instance, a study showed acute coronary syndrome (ACS) patients who took part in CR exhibited significant increases in QOL scores across all domains [16]. This improvement in QOL is attributed to the comprehensive nature of CR, which addresses not only physical health but also psychological and social aspects.

Yoga-based Cardiac Rehabilitation led to enhanced self-assessed well-being and a return to activities as they were before the heart attack, following an acute myocardial infarction. However, the study did not have enough statistical strength to demonstrate a distinction in major adverse cardiovascular events (MACE). In cases where traditional cardiac rehabilitation is inaccessible or not preferred by individuals, Yoga-CaRe could present itself as a potential alternative [17].

Due to these numerous favourable effects, it is essential to promote greater participation and referral of cardiac rehabilitation (CR) in numerous countries globally. This is particularly important since many nations experiencing a high incidence of ischemic heart disease possess a limited quantity of CR programs.

Despite the substantial evidence supporting the benefits of CR, several limitations should be acknowledged. Variability in the design of CR programs, differences in patient populations, and varying lengths of follow-up periods can introduce heterogeneity in study outcomes. Moreover, some studies have failed to show statistically significant differences in certain outcomes. Future research should aim to standardize CR interventions and implement rigorous methodologies to enhance the robustness of findings.

## Conclusion

Cardiac rehabilitation is an integral component of cardiovascular care, recommended by clinical guidelines to improve patient outcomes and their quality of life. By addressing both physical and psychological aspects, CR contributes to the reduction of cardiovascular mortality, hospital admissions, and adverse events. The lack of statistical evidence underpowers the value of CR in patient well-being and long-term cardiovascular health. Continued research and efforts to promote the widespread adoption of CR programs are crucial to ensuring better outcomes for patients with myocardial infarction.

## Limitations

Our literature review has limitations. We limited our analysis to English articles published within the last 10 years, specifically targeting those at least 19 years old. We also only used free articles, and our study was limited to English papers on Cardiac rehabilitation in Post Myocardial Infarction. More research is needed for specific conclusions.

## Data Availability

All data produced in the present work are contained in the manuscript

## Abbreviations used

(MI): Myocardial Infarction
(AMI): Acute myocardial infarction
(ACS): Acute Coronary Syndrome
(CVDs): Cardiovascular diseases
(CR): Cardiac rehabilitation
(QOL): Improved quality of life

